# Lung function may recover after coal mine fire smoke exposure: a longitudinal cohort study

**DOI:** 10.1101/2024.07.29.24311157

**Authors:** Nicolette R Holt, Catherine L Smith, Caroline X Gao, Brigitte Borg, Tyler J Lane, David Brown, Jillian F Ikin, Annie Makar, Thomas McCrabb, Mikayla Thomas, Kris Nilsen, Bruce R Thompson, Michael J Abramson

## Abstract

**Background and objective:** The 2014 Hazelwood coalmine fire exposed residents in nearby Morwell to high concentrations of particulate matter <2·5 µm (PM_2·5_) for approximately 6 weeks. This analysis aimed to evaluate the long-term impact on respiratory health.

**Methods:** Adults from Morwell and the unexposed town of Sale completed validated respiratory questionnaires and performed spirometry, gas transfer and oscillometry 3·5-4 years (Round 1) and 7·3-7·8 years (Round 2) after the fire. Individual PM_2·5_ exposure levels were estimated using chemical transport models mapped onto participant-reported time-location data. Mixed-effects regression models were fitted to analyse associations between PM_2·5_ exposure and outcomes, controlling for key confounders.

**Results:** From 519 (346 exposed) Round 1 participants, 329 (217 exposed) participated in Round 2. Spirometry and gas transfer in Round 2 were mostly lower compared with Round 1, excepting FVC (increased) and FEV_1_ (minimal change). The effect of mine fire-related PM_2·5_ exposure changed from a negative effect in Round 1 to no effect in Round 2 for both pre-(p=0·005) and post-bronchodilator FVC (p=0·032). PM_2·5_ was not associated with gas transfer in either round. For post-bronchodilator reactance and area under the curve, a negative impact of PM_2·5_ in Round 1 showed signs of recovery in Round 2 (both p<0·001).

**Conclusion:** In this novel study evaluating long-term respiratory outcomes after medium-duration high concentration PM_2·5_ exposure, the attenuated associations between exposure and respiratory function may indicate some recovery in lung function. With increased frequency and severity of landscape fires observed globally, these results inform public health policies and planning.

**KEY MESSAGES:** Evidence is currently lacking on the long-term sequelae of high concentration PM_2·5_ exposure, from extreme wildfire events lasting weeks to months, on lung physiology and function. We found that previously observed deficits in adult lung function, measured using spirometry, gas transfer and oscillometry 3.5 years after a prolonged coal mine fire, may recover in the longer-term. With increased frequency of prolonged landscape fires observed globally, these results inform public health policies and planning.

## INTRODUCTION

Continued exposure to ambient air pollution, especially particulate matter (PM) from sources such as industry, vehicle exhaust, biomass fuels, and wildfires, is leading to premature deaths and disease worldwide. The Global Burden of Disease study 2019 highlighted air pollution as a leading contributor to global disease burden and premature deaths, a burden disproportionately borne by low-and middle-income countries.[1] In particular, fine PM with a median aerodynamic diameter < 2·5 µm (PM_2·5_) is able to infiltrate deep into the lung periphery and the blood stream.[2, 3] It was estimated that 4·1 million premature deaths in 2019, representing 7·3% of total deaths globally and 4·7% of global disability-adjusted life-years (DALYs), were attributable to the long-term exposures to PM_2·5._[4, 5] There is a strong causal association between PM_2·5_ exposure and cardiopulmonary diseases.[6] Increasing PM_2·5_ exposure and chronic obstructive pulmonary disease (COPD) prevalence and lung function decline have been well documented.[7] Evidence is currently lacking on the long-term sequelae of PM_2·5_ exposure from medium-term ambient PM_2·5_ exposure (weeks to months) from extreme events such as wildfires on lung physiology and function.[8] Indeed, studies of wildfires often utilise secondary data, such as hospital presentation records, to infer respiratory associations.[9] With climate change, and the increasing frequency as well as intensity of wildfires globally, addressing these gaps in evidence is critical to formulation of health policies.[10]

In February 2014, a fire in the Hazelwood open-cut brown coal mine, located in the Latrobe Valley, south-eastern Australia, exposed nearby residents to significant ambient air pollution for approximately 6 weeks. The adjacent town of Morwell, with a population at the time numbering approximately 14,000, experienced the most extreme smoke levels. Hourly mine fire-related PM_2.5_ concentrations were estimated to have reached 3700 µg/m^3^ [11, 12] during the initial phase of the fire. The daily average National Environment Protection Measure (NEPM) standard of 25 µg/m^3^ was breached on 27 days during February and March 2014 in Morwell.[12] A Victorian Government appointed Board of Inquiry into the mine fire heard that the Latrobe Valley community, particularly those in Morwell, reported a variety of physical symptoms including sore and stinging eyes, coughing, shortness of breath, headaches, chest pain, fatigue, mouth ulcers, blood noses and rashes.[13]

The Hazelwood Health Study (HHS; hazelwoodhealthstudy.org.au) was established to investigate the long-term adverse health outcomes in people exposed to the mine fire smoke. The Study’s Hazelinks Stream, which analysed administrative health service use data, reported dose response associations between increasing PM_2·5_ exposure and increased respiratory-related ambulance attendances,[14] emergency department presentations and hospital admissions,[15] General Practitioner (GP) and specialist consultations[16] and dispensing of medications.[17]

The HHS Adult Cohort (N=4056) was established in 2016 and comprised adult residents of Morwell (exposed) and Sale (unexposed but otherwise similar town – refer to Table 1 in Holt et al[18]) who completed the Adult Survey.[19, 20] A subgroup of 519 cohort members subsequently participated in Round 1 (R1) of the HHS adult Respiratory Stream in 2017-2018.[18] Adult Cohort members who had reported an asthma attack or current asthma medication use in the Adult Survey were oversampled (40%) to provide ability for further evaluation of effects in asthmatics. The HHS adult Respiratory Stream aimed to investigate the association between exposure to mine fire-related PM_2·5_ and lung function at approximately 3, 6 and 9 years after the event.[18] At R1, 3.5 years after the fire, there was a clear dose response relationship between medium-duration, but extreme mine fire-related PM_2·5_ exposure and spirometry consistent with COPD in non-smokers[21] and worsening lung mechanics.[18]

In a second round of respiratory testing (R2) conducted in 2021, we aimed to investigate the longer-term impacts of mine fire-related PM_2·5_ exposure on respiratory health. Because background ambient PM_2·5_ exposure had the potential to confound associations between fire-related PM_2·5_ exposure and health, and because many of the HHS participants were potentially exposed to ambient smoke from the 2019-20 ‘Black Summer’ bushfires (wildfires), examination of PM_2·5_ levels during these bushfires was conducted.

## METHODS

### Study design and setting

The Respiratory Stream of the HHS is a longitudinal cohort study. A flowchart showing the study recruitment process is provided in Figure 1. Recruitment for R2 was originally planned for 2020, however that was postponed due to COVID-19-related lockdowns. R2 data collection went ahead in the Latrobe Valley between May and November 2021. The study was conducted in compliance with the principles of the Declaration of Helsinki. The protocol and amendments were approved by the Monash University and Alfred Health Ethics Committees. All participants provided written informed consent.

### Public involvement

The Latrobe Valley community has had ongoing involvement in the design and conduct of the HHS. In the immediate aftermath of the mine fire, a Board of Inquiry into the Hazelwood coalmine fire held ten community consultations encouraging members to describe their experiences and concerns. The Board also received more than 700 public submissions and heard from six independent experts and 13 community witnesses. The HHS was subsequently designed in direct response to the community’s concerns. The HHS Community Advisory Committee (CAC) was later convened in February 2015 and has met at least quarterly since. The CAC initially comprised five community lay persons and representatives from seven local organisations (Latrobe City Council, Latrobe Community Health Service, Latrobe Regional Hospital, the Department of Health Gippsland, Federation University, Wellington Shire and the Central Gippsland Health Service Board). In July 2021, the CAC merged with the activities of the existing Latrobe Health Assembly (LHA) and became the LHA HHS sub-committee, comprising four LHA members and four community members. The Committee provides advice on all aspects of the HHS’s activities including study design, recruitment and disseminations of findings.

### Participant recruitment and characteristics

Participants were eligible for the 2021 Respiratory Stream R2 assessment if they had participated in R1 in 2017.[18] Participants were excluded if they were aged over 90 years, or identified to be deceased. Otherwise eligible participants were also excluded if a contraindication to spirometry was identified – including recent surgery, myocardial infarction, pneumothorax, pulmonary embolism, open pulmonary tuberculosis or known aneurysms.[22] Recruitment was via mailed invitation, email, Short Message Service or phone call, depending on last known contact details. Measurement of participant characteristics varied across the Adult Survey and clinical assessment rounds. Level of education, ethnicity, sex, and town (Morwell/Sale) were obtained from the Adult Survey.[20] In both R1 and R2, height and weight were measured during the clinic assessments, and age, employment, and smoking history collected via questionnaires. Participants were classified as current, ex-(current non-smokers with >100 lifetime cigarettes) or non-smokers (<100 lifetime cigarettes).[23]

### Exposure assessment

Individual level mine fire exposure for all participants was previously calculated by mapping the spatial and temporal distribution of mine fire-related PM_2·5_ concentrations, which had been estimated using chemical transport models, onto participants’ time-location data captured as a part of the Adult Survey; detailed elsewhere.[18–20] Mean 24-hour mine fire-related PM_2·5_ concentrations experienced by participants ranged from 0 to 56 µg/m^3^. In addition to the coalmine fire, participants were also potentially exposed to smoke from the 2019-20 ‘Black Summer’ bushfires, with Sale (unexposed to the mine fire), being closer to the region with major fires (far eastern Victoria) than Morwell. We previously utilised PM_2·5_ data from the Environment Protection Authority (EPA) of Victoria to evaluate possible exposure variations between Morwell and Sale, as a result of this event. While ambient PM_2·5_ has the potential to confound associations, we found little difference in annual mean ambient PM_2·5_ between Morwell (7.9 μg/m^3^) and Sale (7.6 μg/m^3^) from 2009 to 2022[24, 25] which included the bushfire period.

### Clinical measures

Respiratory testing was performed by trained respiratory scientists at all sites for both data collection rounds, following standard operating procedures and in accordance with current respiratory measurement standards where available. Spirometry and gas transfer (transfer or diffusing factor of the lung for carbon monoxide; TLCO) were measured using the EasyOne Pro Lab Respiratory Analysis System (ndd Medical Technologies AG, Zürich, Switzerland) in line with international standards.[26–28]

Oscillometry was assessed using the Forced Oscillation Technique (FOT) with the Tremoflo C-100 device (Thorasys, Montreal, Canada) in line with standards current at time of testing.[29, 30] Indices reported included resistance at 5Hz (R5), difference in resistance at 5 and 19Hz (R5-19), reactance at 5Hz (X5), and the area under the reactance curve at 5Hz (Ax5). Test acceptability was evaluated using guidelines on coherence criteria available at the time of the test.

Spirometry and oscillometry were performed before and after administration of short-acting bronchodilator (300µg salbutamol). Bronchodilator use in the previous 24 hours was recorded, as bronchodilator therapy was unable to be withheld prior to assessment due to ethical reasons.

Participants were considered as having COPD if spirometry demonstrated a post-bronchodilator (BD) ratio of FEV_1_ to forced vital capacity (FEV_1_/FVC) < lower limit of normal (fifth percentile) using the Global Lung Initiative (GLI) spirometry reference values.[31] Self-reported asthma was captured via a modified European Community Respiratory Health Survey questionnaire[32] in both rounds.

Spirometry quality was assessed according to ATS/ERS technical standards applicable at time of testing and graded accordingly.[27, 28, 33] Results graded less than A or B were reviewed by an experienced, senior respiratory scientist (BB) and included or excluded based on likely validity of results.

### Statistical analysis

Descriptive statistics (such as means and standard deviations [SD]) were used to report characteristics and outcome differences between rounds. Mean differences in outcomes between the two rounds were compared to a mean of zero using one sample t-tests. To investigate potential selection bias, participant characteristics and outcomes at R1 were compared between the participants and non-participants of the R2 testing.

Z-scores of spirometry and gas transfer indices, calculated from the Global Lung Function Initiative reference equations for Spirometry and TLCO,[31, 34] were used as outcome variables. Non-linear transformations were used for oscillometry variables, as a high proportion of our participants fell outside the required prediction range for the published algorithm[18, 35] with z-scores used in sensitivity analyses.

To model the effect of mine fire-related PM_2·5_ exposure on each outcome over time, linear mixed effects models with random intercepts were used to account for repeated measurements. The models measured the effects of: (1) mine-fire related PM_2·5_ exposure (10µg/m^3^ increase), (2) changes over time (from R1 to R2 using an indicator variable) and (3) whether the effect of exposure on the outcome changed over time (interaction between exposure and time). To assist with interpretation, the results were reported as effects of exposure (coefficients and 95% Confidence Intervals [CI]) on the outcome at R1 and R2 using linear combination of coefficients as well as p-values for interaction terms (indicating whether the two coefficients were statistically distinguishable).

All models controlled for potential confounding factors (including education and employment, smoking status, self-reported asthma, spirometric COPD and other factors related to testing: age, gender, height, and weight were included for outcomes without z-scores) and were weighted to correct for oversampling of asthmatics. Pre bronchodilator outcomes were also adjusted for whether inhaled medication was withheld by asthmatics prior to spirometry. Multiple imputation was used to account for missing data in all regression analyses.[36] Data were analysed using Stata v16 (StataCorp, 2College Station, TX 2017).

## RESULTS

### Participant characteristics

From 519 (346 exposed from Morwell, 173 from Sale) R1 participants, 329 (217 exposed) participated in R2. **Figure 1** shows the flow chart of participants. **Table 1** presents participant characteristics by respiratory assessment round. Participant demographic characteristics were comparable across clinical assessment rounds, except for higher loss to follow-up among current smokers. **Table S1** in Supporting Information provides full details. Two-thirds of participants were from Morwell, and 59% were female. The mean age was 54.9 years (SD 16.1) in R1 and 57.9 years (SD 15.2) in R2. Half the participants were categorised as obese (Body Mass Index; BMI≥30kg/m^2^), around 45% reported an asthma diagnosis, and approximately 13% had COPD based on spirometry at R1.

**Figure 1.**
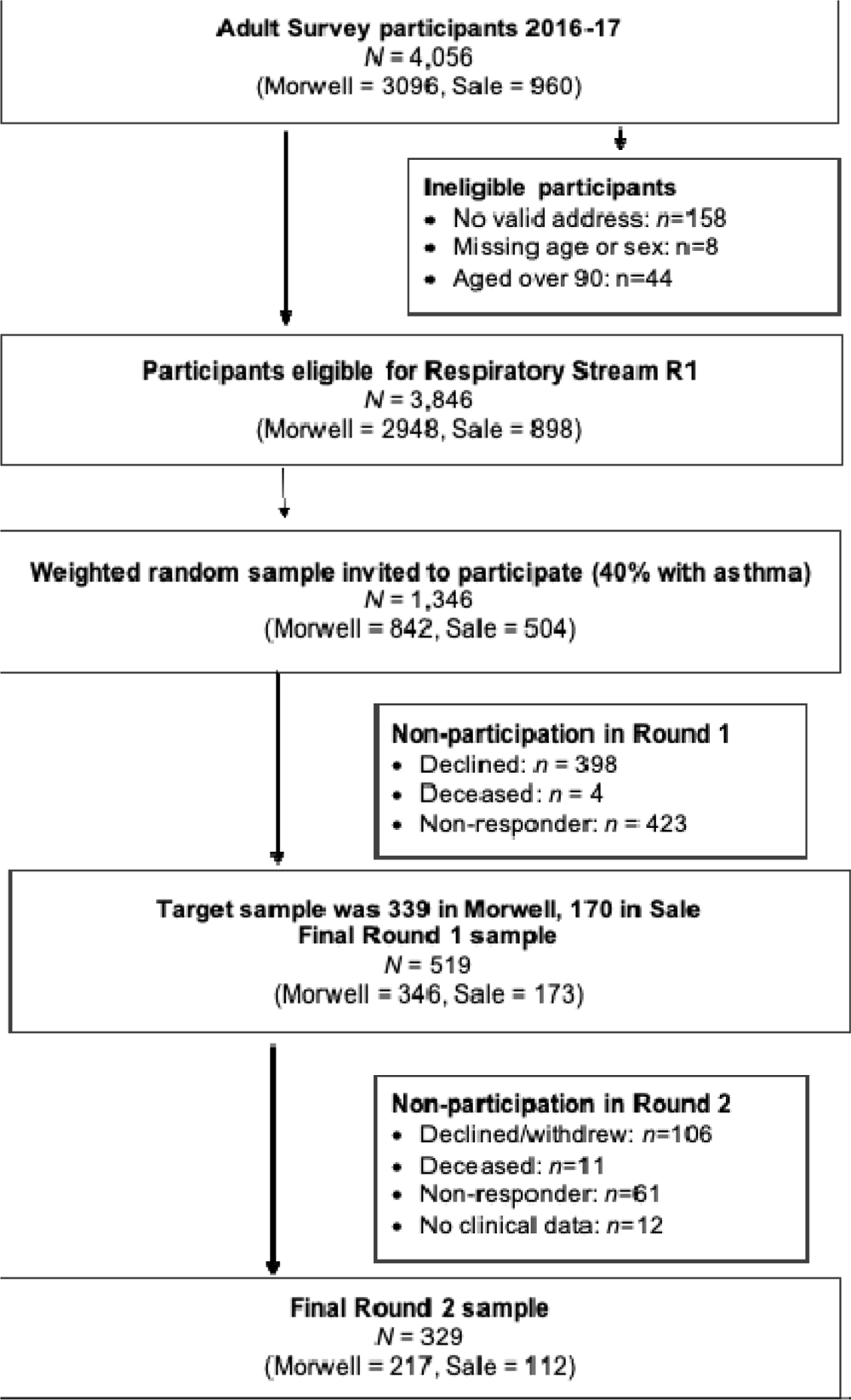
Hazelwood Health Study Respiratory Stream recruitment

**Table 1:** Participant characteristics at the time of each clinical assessment round.

| Characteristic | Round 1<br>N=519 | Round 2<br>N=329 |
| --- | --- | --- |
| **Town, n (%)** |  |  |
| Morwell | 346 (67%) | 217 (66%) |
| Sale | 173 (33%) | 112 (34%) |
| **At the time of the mine fire: Daily exposure to fire-related PM<sub>2.5</sub> (µg/m<sup>3</sup>), median [Q1-Q3]** |  |  |
| Morwell | 11.7 [7.2-18.4] | 11.8 [7.2-17.4] |
| Sale | 0.0 [0.0-0.0] | 0.0 [0.0-0.0] |
| **Age, mean (SD)** | 54.9 (16.1) | 57.9 (15.2) |
| **Age category at each round, n (%)** |  |  |
| < 45 years | 151 (29%) | 73 (22%) |
| 45-65 years | 211 (41%) | 139 (42%) |
| > 65 years | 157 (30%) | 117 (36%) |
| **Female, n (%)** | 306 (59%) | 195 (59%) |
| **Employment status, n (%)** |  |  |
| Employed | 229 (44%) | 143 (43%) |
| Other (retired, home duties, study, other) | 216 (42%) | 153 (47%) |
| Unemployed / unable to work | 74 (14%) | 33 (10%) |
| **Post-secondary education, n (%)** | 291 (57%) | 196 (60%) |
| **Height (cm), mean (SD)** | 166 (9) | 167 (9) |
| **Weight (kg), mean (SD)** | 85.6 (21.3) | 86.6 (21.2) |
| **BMI (kg/m<sup>2</sup>), mean (SD)** | 31.0 (7.4) | 30.8 (7.4) |
| **BMI, n (%)** |  |  |
| Underweight / Normal (BMI<25 kg/m<sup>2</sup>) | 99 (19%) | 66 (20%) |
| Overweight (25≤BMI<30 kg/m<sup>2</sup>) | 167 (32%) | 100 (30%) |
| Obese (BMI≥30 kg/m<sup>2</sup>) | 253 (49%) | 163 (50%) |
| **Self-reported Asthma, n (%)** | 229 (44%) | 147 (45%) |
| **Spirometric COPD, n (%)** | 59 (12%) | 41 (13%) |
| **Smoking status, n (%)** |  |  |
| Non-smoker | 249 (48%) | 173 (53%) |
| Ex-smoker | 186 (36%) | 114 (35%) |
| Current smoker | 84 (16%) | 42 (13%) |

Abbreviations: BMI = body mass index, COPD = chronic obstructive pulmonary disease, SD = Standard Deviation, PM<sub>2.5</sub> = particulate matter <2.5 µg/m<sup>3</sup>, Q1 = first quartile, Q3 = third quartile.
Missing: education *n* = 10; height *n* = 2; weight *n* = 3; BMI *n* = 3; COPD *n* = 20

Examination of PM_2·5_ levels during the 2019-20 ‘Black Summer’ bushfires suggested no evidence of different exposures between the two towns of Morwell and Sale (refer to Appendix S1 and Figures S1 and S2 in Supporting Information).

Descriptive statistics for the study outcomes (z-scores for spirometry and gas transfer; transformed scores for oscillometry) are given in **Table 2** with crude values provided in **Table S3** in Supporting Information and z-scores for oscillometry variables in **Table S4** in Supporting Information. Spirometry and gas transfer z-scores in R2 were slightly lower compared with R1 assessment, except for FVC.

Spirometry quality was very good with less than 1% of R1 tests, and only 1.5% of R2 tests, excluded based on likely validity of results (**Table S5** in Supporting Information).

**Table 2:**
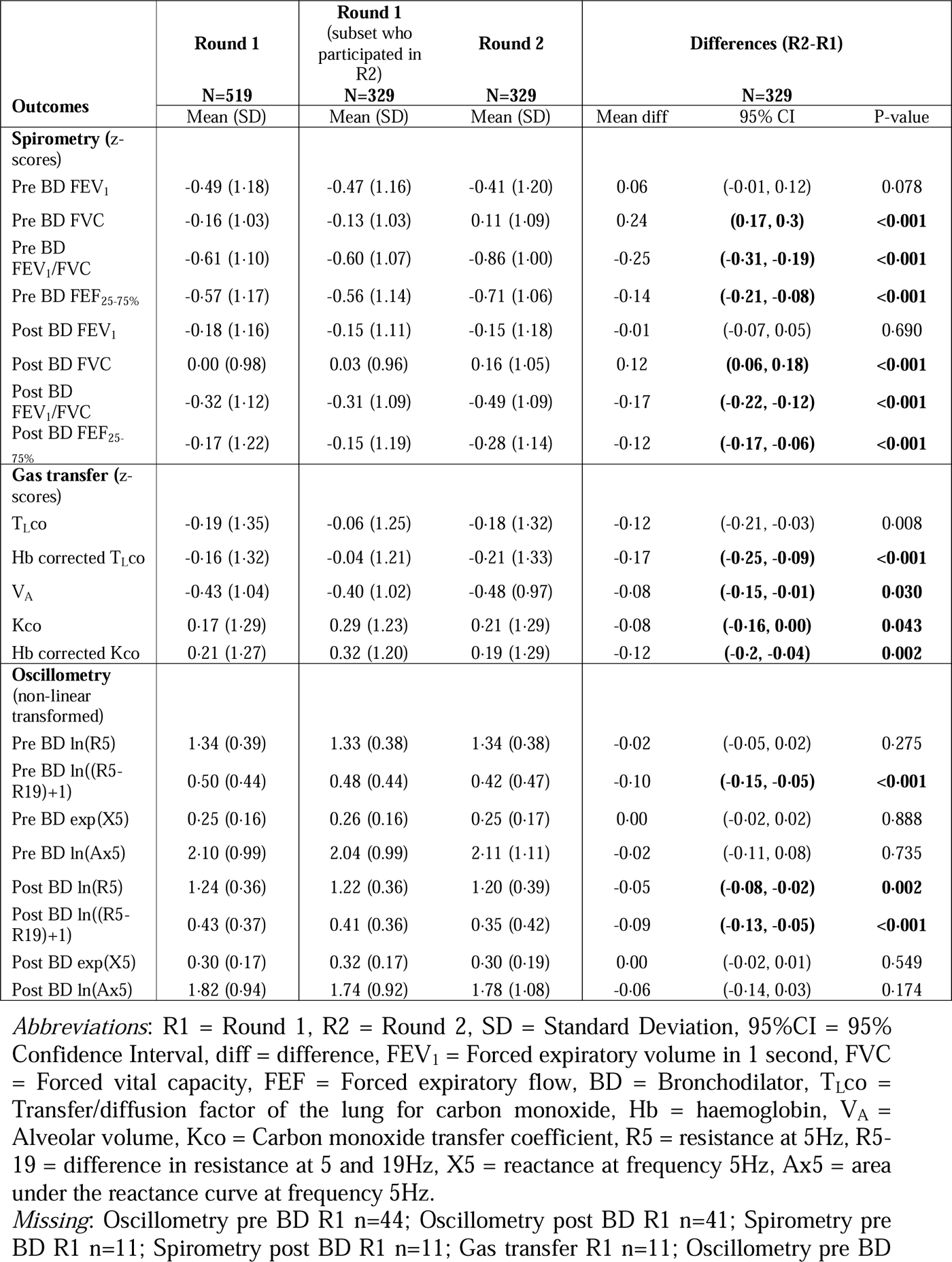
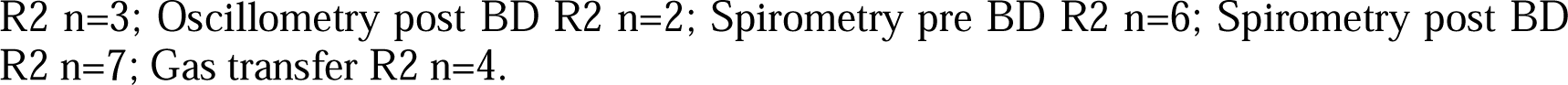
Respiratory outcome means (z-scores or transformed) by assessment round and crude mean differences for those completing both rounds (R2-R1)

| Outcomes | Round 1 | Round 1<br>(subset who<br>participated in<br>R2) | Round 2 | Differences (R2-R1) |  |  |
| --- | --- | --- | --- | --- | --- | --- |
|  | N=519 | N=329 | N=329 | N=329 |  |  |
|  | Mean (SD) | Mean (SD) | Mean (SD) | Mean diff | 95% CI | P-value |
| **Spirometry** (z-scores) |  |  |  |  |  |  |
| Pre BD FEV<sub>1</sub> | -0.49 (1.18) | -0.47 (1.16) | -0.41 (1.20) | 0.06 | (-0.01, 0.12) | 0.078 |
| Pre BD FVC | -0.16 (1.03) | -0.13 (1.03) | 0.11 (1.09) | 0.24 | **(0.17, 0.3)** | **<0.001** |
| Pre BD FEV<sub>1</sub>/FVC | -0.61 (1.10) | -0.60 (1.07) | -0.86 (1.00) | -0.25 | **(-0.31, -0.19)** | **<0.001** |
| Pre BD FEF<sub>25-75%</sub> | -0.57 (1.17) | -0.56 (1.14) | -0.71 (1.06) | -0.14 | **(-0.21, -0.08)** | **<0.001** |
| Post BD FEV<sub>1</sub> | -0.18 (1.16) | -0.15 (1.11) | -0.15 (1.18) | -0.01 | (-0.07, 0.05) | 0.690 |
| Post BD FVC | 0.00 (0.98) | 0.03 (0.96) | 0.16 (1.05) | 0.12 | **(0.06, 0.18)** | **<0.001** |
| Post BD FEV<sub>1</sub>/FVC | -0.32 (1.12) | -0.31 (1.09) | -0.49 (1.09) | -0.17 | **(-0.22, -0.12)** | **<0.001** |
| Post BD FEF<sub>25-75%</sub> | -0.17 (1.22) | -0.15 (1.19) | -0.28 (1.14) | -0.12 | **(-0.17, -0.06)** | **<0.001** |
| **Gas transfer** (z-scores) |  |  |  |  |  |  |
| T<sub>L</sub>co | -0.19 (1.35) | -0.06 (1.25) | -0.18 (1.32) | -0.12 | (-0.21, -0.03) | 0.008 |
| Hb corrected T<sub>L</sub>co | -0.16 (1.32) | -0.04 (1.21) | -0.21 (1.33) | -0.17 | **(-0.25, -0.09)** | **<0.001** |
| V<sub>A</sub> | -0.43 (1.04) | -0.40 (1.02) | -0.48 (0.97) | -0.08 | **(-0.15, -0.01)** | **0.030** |
| Kco | 0.17 (1.29) | 0.29 (1.23) | 0.21 (1.29) | -0.08 | **(-0.16, 0.00)** | **0.043** |
| Hb corrected Kco | 0.21 (1.27) | 0.32 (1.20) | 0.19 (1.29) | -0.12 | **(-0.2, -0.04)** | **0.002** |
| **Oscillometry** (non-linear transformed) |  |  |  |  |  |  |
| Pre BD ln(R5) | 1.34 (0.39) | 1.33 (0.38) | 1.34 (0.38) | -0.02 | (-0.05, 0.02) | 0.275 |
| Pre BD ln((R5-R19)+1) | 0.50 (0.44) | 0.48 (0.44) | 0.42 (0.47) | -0.10 | **(-0.15, -0.05)** | **<0.001** |
| Pre BD exp(X5) | 0.25 (0.16) | 0.26 (0.16) | 0.25 (0.17) | 0.00 | (-0.02, 0.02) | 0.888 |
| Pre BD ln(Ax5) | 2.10 (0.99) | 2.04 (0.99) | 2.11 (1.11) | -0.02 | (-0.11, 0.08) | 0.735 |
| Post BD ln(R5) | 1.24 (0.36) | 1.22 (0.36) | 1.20 (0.39) | -0.05 | **(-0.08, -0.02)** | **0.002** |
| Post BD ln((R5-R19)+1) | 0.43 (0.37) | 0.41 (0.36) | 0.35 (0.42) | -0.09 | **(-0.13, -0.05)** | **<0.001** |
| Post BD exp(X5) | 0.30 (0.17) | 0.32 (0.17) | 0.30 (0.19) | 0.00 | (-0.02, 0.01) | 0.549 |
| Post BD ln(Ax5) | 1.82 (0.94) | 1.74 (0.92) | 1.78 (1.08) | -0.06 | (-0.14, 0.03) | 0.174 |

*Abbreviations:* R1 = Round 1, R2 = Round 2, SD = Standard Deviation, 95%CI = 95% Confidence Interval, diff = difference, FEV<sub>1</sub> = Forced expiratory volume in 1 second, FVC = Forced vital capacity, FEF = Forced expiratory flow, BD = Bronchodilator, T<sub>L</sub>co = Transfer/diffusion factor of the lung for carbon monoxide, Hb = haemoglobin, V<sub>A</sub> = Alveolar volume, Kco = Carbon monoxide transfer coefficient, R5 = resistance at 5Hz, R5-19 = difference in resistance at 5 and 19Hz, X5 = reactance at frequency 5Hz, Ax5 = area under the reactance curve at frequency 5Hz.
*Missing:* Oscillometry pre BD R1 n=44; Oscillometry post BD R1 n=41; Spirometry pre BD R1 n=11; Spirometry post BD R1 n=11; Gas transfer R1 n=11; Oscillometry pre BD
R2 n=3; Oscillometry post BD R2 n=2; Spirometry pre BD R2 n=6; Spirometry post BD R2 n=7; Gas transfer R2 n=4.

### The associations between mine fire-related PM_2·5_ exposure and lung function

Figure 2 shows the estimated effect of mine fire-related PM_2·5_ exposure on each lung function outcome at R1 and R2 as well as the p-values for the interaction term between exposure and Round (p_int_).

**Figure 2a.**
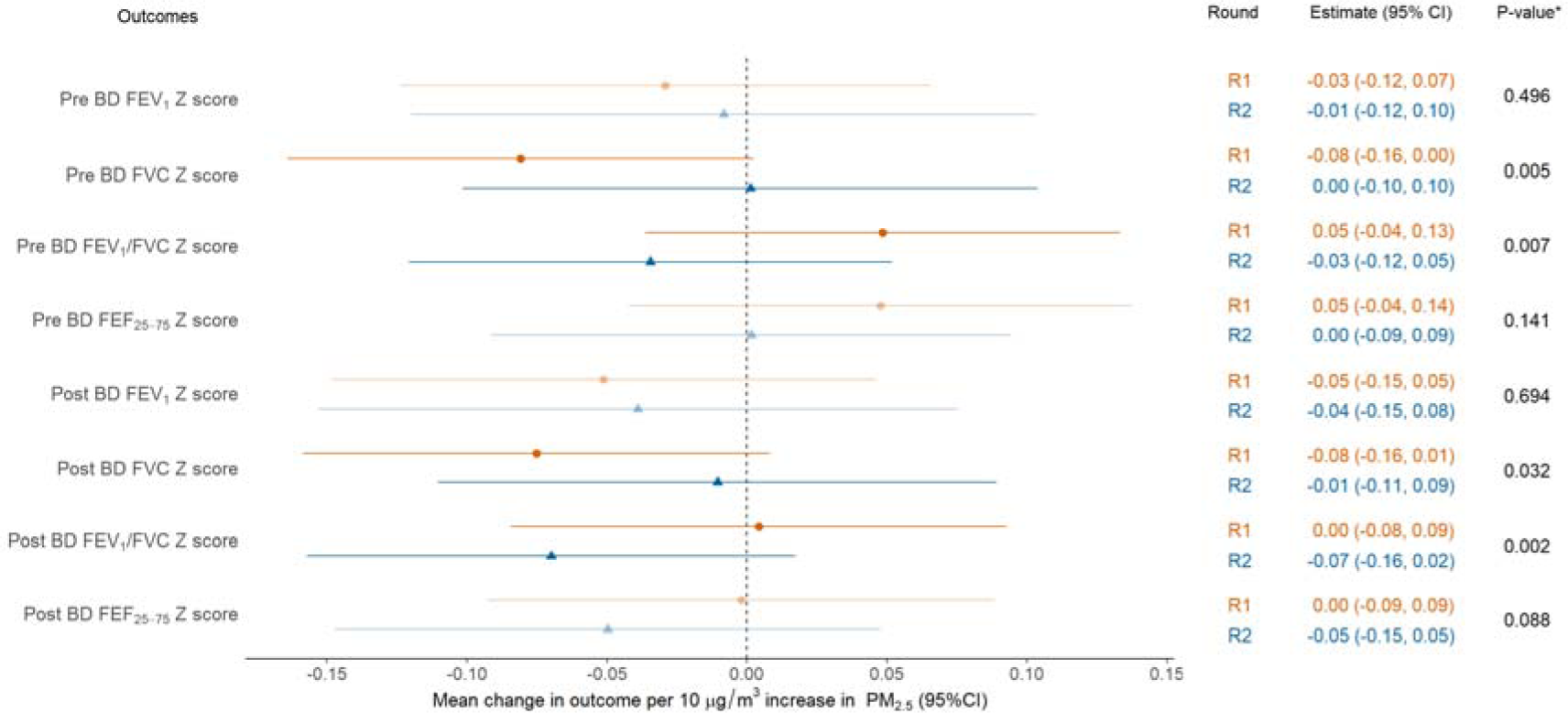
Models of Spirometry as a function of mine fire-related PM_2·5_ * p-values for the interaction term between exposure and Round. All models adjusted for education, employment, asthma, and smoking status. Pre BD outcomes also adjusted for whether inhaled medication was withheld. *Abbreviations*: R1 = Round 1, R2 = Round 2, PM_2·5_ = particulate matter <2·5 µg/m³, 95%CI = 95% Confidence Interval, FEV_1_ = Forced expiratory volume in 1 second, FVC = Forced vital capacity, FEF = Forced expiratory flow, BD = Bronchodilator.

**Figure 2b.**
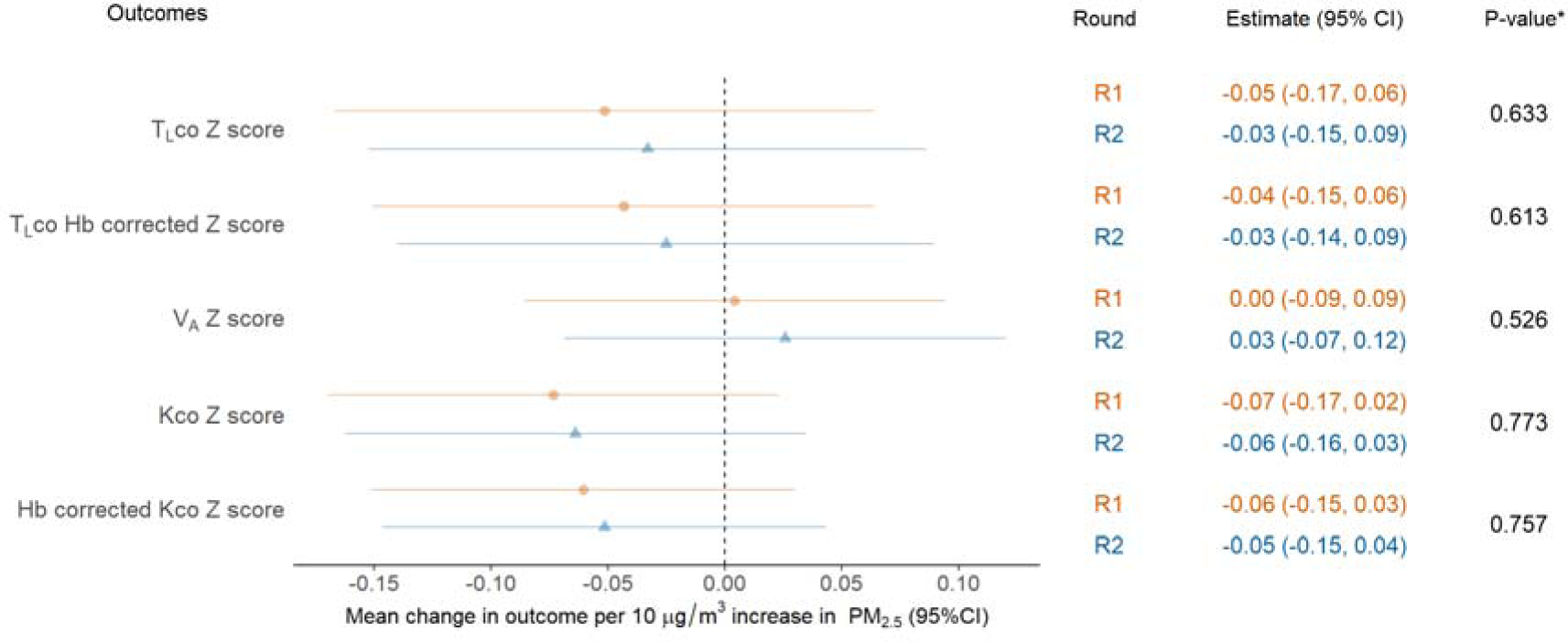
Models of gas transfer as a function of mine fire-related PM_2·5_ *p-values for the interaction term between exposure and Round. All models adjusted for education, employment, spirometric COPD and smoking status.

**Figure 2c.**
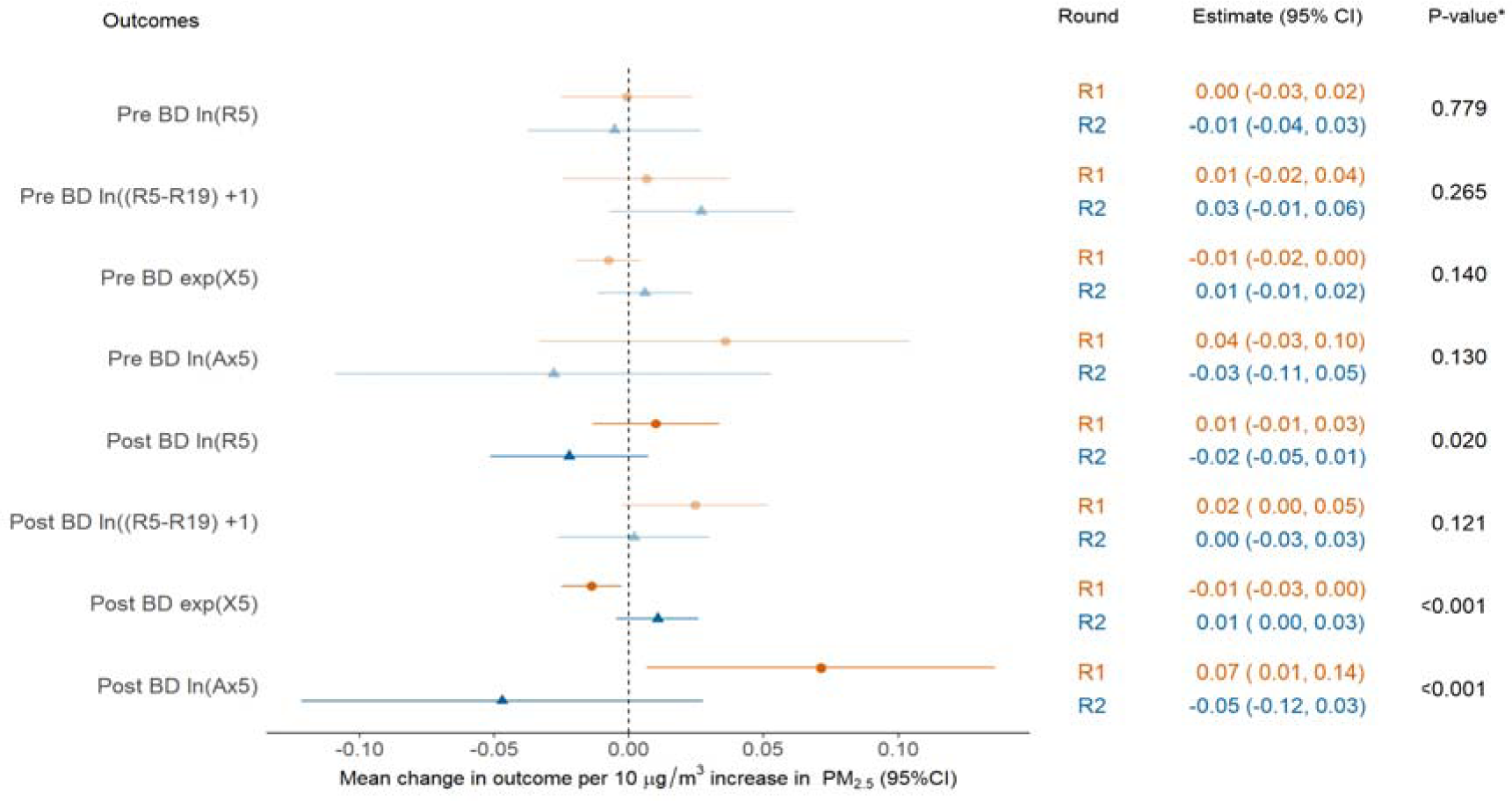
Models of oscillometry as a function of mine fire-related PM_2.5_ *p-values for the interaction term between exposure and Round. All models were adjusted for age, gender, height, weight, education, employment, asthma, spirometric COPD and smoking status. Pre BD outcome also adjusted for whether inhaled medication was withheld prior to spirometry. *Abbreviations:* R1 = Round 1, R2 = Round 2, PM_2·5_ = particulate matter <2·5 µg/m³, SD = Standard Deviation, 95%CI = 95% Confidence Interval, BD = Bronchodilator, R5 = resistance at 5Hz, R5-19 = difference in resistance at 5 and 19Hz, X5 = reactance at 5Hz, Ax5 = area under the reactance curve at 5Hz.

Spirometry and gas transfer in R2 were lower compared with R1, excepting FVC (unadjusted mean increase 60mL). Using FVC z-score as an example (Figure 2A), there was a small mean decrease of 0·08 in baseline (pre BD) FVC z-score for every 10µg/m^3^ increase in mine fire-related PM_2·5_ exposure (95%CI: −0·16 to 0·00) in R1, which was reduced to no mean change of 0·0 (−0·10 to 0·11) in R2. The overall effect of exposure changed over time from a negative effect in R1 to no effect in R2 (p_int_=0·005). This effect was similar to Post BD FVC z-score (p_int_=0·032). As a result, a reversed direction of association for FEV_1_/FVC z-scores was evident both at pre BD (p_int_=0·007) and post BD (p_int_=0·002). However, generally, the estimated mean change in spirometry outcomes with increasing PM_2·5_ exposure overlapped with the null effect in both rounds (Figure 2A). Similarly, exposure to mine fire-related PM_2·5_ was not found to be associated with gas transfer at either round, and the effect was unchanged between rounds (Figure 2B).

Figure 2C shows the estimated effect of PM_2·5_ exposure on transformed oscillometry outcomes. The main changes were for post BD resistance (ln[R5]), reactance (exp[X5]), and area under the reactance curve (ln[Ax5]). For both post-BD reactance and area under the reactance curve, a negative impact of exposure in R1 showed signs of recovery in R2 (both p_int_<0·001). These results were consistent using the oscillometry z-scores as a function of PM_2·5_ (see **Figure S4** in Supporting Information).

## DISCUSSION

Assessment of participants at 3·5-4·0 (Round 1) and 7·3-7·8 years (Round 2) after the Hazelwood coal mine fire revealed an attenuated association between medium-duration exposure to extreme coal mine fire-related PM_2·5_ and several respiratory function measures. The effect of exposure on spirometry changed over time, from a negative effect in R1 to no effect in R2 for both pre bronchodilator and post-bronchodilator FVC. The estimated mean change in spirometry with increasing PM_2·5_ exposure, overlapped with a null effect in both rounds. This null finding evaluating the association between PM_2·5_ exposure and change in lung function is consistent with prior community-based cohort studies from Europe.[37, 38] This was the first study utilising oscillometry to evaluate the long-term respiratory impact after medium-duration coal mine fire smoke-related PM_2·5_ exposure. R1 revealed a clear dose-response association between coal mine fire-related PM_2·5_ exposure and a more negative respiratory system reactance.[18] We acknowledge that part of this observed effect may be attributable to normal aging. The underlying mechanism remains unclear, although one possible explanation might be early peripheral airway changes with accelerated pulmonary ageing. For the estimated effect of PM_2·5_ exposure on transformed oscillometry outcomes, both post-bronchodilator reactance and area under the reactance curve, a negative impact of exposure in R1 showed signs of recovery in R2. This observation may reflect potential recovery of lung function following an episode of medium-duration high intensity PM_2·5_ exposure.

Oscillometry assesses lung mechanics and may detect of early changes in peripheral airway function that conventional spirometry cannot.[29] Oscillometry has previously been used to evaluate changes in lung mechanics in firefighters exposed to asbestos[39] and dust exposure from the World Trade Centre following the 9/11 attacks,[40, 41] where spirometry was normal, but abnormalities were detected in oscillometry. To our knowledge, no prior study has evaluated the long-term sequelae of PM_2·5_ exposure from landscape fires, or biomass fuel smoke using oscillometry. Our findings add valuable insights into the impact of coal mine fire-related PM_2.5_ on lung function.

Wildfire particulate matter may be more harmful than equivalent exposures to urban background PM [42] This could also apply to coal mine fire PM . The World Health Organization (WHO) Global Air Quality Guidelines (AQGs) provide recommendations on air quality levels and interim targets for six key air pollutants.[43] However, they do not take into account variations in PM_2·5_ toxicity from different emission sources or the pattern or exposure, such as acute high-intensity compared to chronic exposure. Assessing the relative impact of PM_2·5_ exposure from landscape fire smoke relative to other PM_2·5_ exposure, such as exhaust emissions, is a pressing public health concern, particularly in the face of global warming and increased frequency and severity of landscape fires. Public health policy should consider the different toxicity of PM_2·5_ emission sources when formulating recommendations and guidelines and planning appropriate public health responses for future episodes.

### Strengths and limitations

This study has many strengths. The sample was drawn from a population based survey. This research has utilised various objective measures of lung function (spirometry, gas transfer and oscillometry) to evaluate respiratory health. To date, most observational studies have typically relied upon secondary data sets eg. hospitalisations.[42] With a second round of data collection, we were able to conduct a longitudinal analysis of the impact of PM_2·5_ exposure on changes in lung function. The models adjusted for relevant confounders, including smoking. A further strength was use of individual mine fire-related PM_2·5_ exposure estimates from chemical transport models and time-location diaries.

Limitations of this study include attrition over the three year follow-up period. The participation rate in Round 2 was affected by movement restrictions associated with the COVID-19 pandemic in Australia. However, nonparticipants were not significantly different from those lost to follow-up, apart from current smoking, which has been frequently observed. It is possible that the reversal of associations with PM_2·5_ could represent loss of statistical power. Wide confidence intervals also indicate statistical uncertainty possibly due to small sample sizes. We did not have individual PM_2·5_ exposure data from the “Black Summer” bushfires nor individual ambient/background PM_2·5_ data. However, examination of ambient PM_2·5_ levels between 2009 and 2022, including during the 2019-20 bushfires, suggested no evidence of different exposures between the two towns of Morwell and Sale.[24] Finally, the analysis conducted multiple comparisons and may contain chance findings.

## Conclusions

The attenuated association between PM_2·5_ exposure and respiratory function measures may indicate some long-term recovery in lung function following medium-term adverse effects. Our study highlighted that oscillometry has an important adjunct role to spirometry in the detection of early changes in lung mechanics associated with environmental and occupational exposures. With climate change driving increased frequency and severity of landscape fires, these results inform public health policies and planning for future events. Further studies are required to confirm the findings and better understand the longer-term respiratory consequences of PM_2·5_ exposure from landscape fire smoke.

## Data Availability

The data are not available as they contain confidential information.

## ABBREVIATIONS

95%CI: 95% Confidence Interval
Ax5: area under the reactance curve at frequency 5Hz.
BD: Bronchodilator
BMI: body mass index
COPD: chronic obstructive pulmonary disease
diff: difference
FEF: Forced expiratory flow
FEV_1_: Forced expiratory volume in 1 second
FVC: Forced vital capacity
Hb: haemoglobin
Kco: Carbon monoxide transfer coefficient
PM_2·5_: particulate matter <2·5 µg/m³
Q1: first quartile
Q3: third quartile
R1: Round 1
R2: Round 2
R5: resistance at 5Hz
R5-19: difference in resistance at 5 and 19Hz
SD: Standard Deviation
T_L_co: Transfer/diffusion factor of the lung for carbon monoxide
V_A_: Alveolar volume
X5: reactance at frequency 5Hz

## ACKNOWLEDGMENTS

The Respiratory Stream clinics were set up in facilities provided by the Central Gippsland Health Service, Sale and The Healthcare Centre, Morwell. We thank Shantelle Allgood and David Poland from Monash Rural Health who oversaw all aspects of participant recruitment, and Sharon Harrison from the Monash School of Public Health and Preventive Medicine for assistance with purchasing, logistics and set up of the clinics.

## DATA AVAILABILITY STATEMENT

Restrictions apply to the availability of these data, which were used under license for this study. Data may be available from the authors but only with the permission of the overseeing ethics committees and the Victorian State Government Department of Health.

