## Supplementary material for "Lung function may recover after coal mine fire smoke exposure: a longitudinal cohort study": NA

### Supporting Information

#### Appendix S1. Black Summer exposure assessment

An initial aim of this exposure assessment was to examine the effects of the 2019-20 “Black Summer”, a bushfire season of unprecedented duration and intensity, and whether it may have confounded analyses by exposing the mine fire-exposed and control sites (Morwell and Sale) to different amounts of smoke. However our analyses of Black Summer data suggested this would not be a major confounder.

The Victorian Environment Protection Authority provided data from the Black Summer, of smoke monitoring sites that contained hourly particulate matter <2·5 µg/m³ (PM_2·5_) measurements. In this analysis, PM_2.5_ (the highest quality standard instrumentation) and iPM_2.5_ (less precise but good for smoke measurements) were used and sPM_2·5_ (sensor network data, much less reliable) excluded. While there were full data for Morwell from two sites (which recorded similar PM_2.5_ levels throughout), there were only a few days of measurements from Sale. Instead, Rosedale, which borders Sale, served as proxy for comparison of PM_2.5_ levels during the Black Summer. There was substantial consistency between the Morwell and Rosedale sites. Notably, peaks in one site corresponded with peaks in the other. However, there were two days in January 2020, which saw the most substantial PM_2.5_ exposure, where the peaks in Rosedale were noticeably higher. This is illustrated in Figure S1.

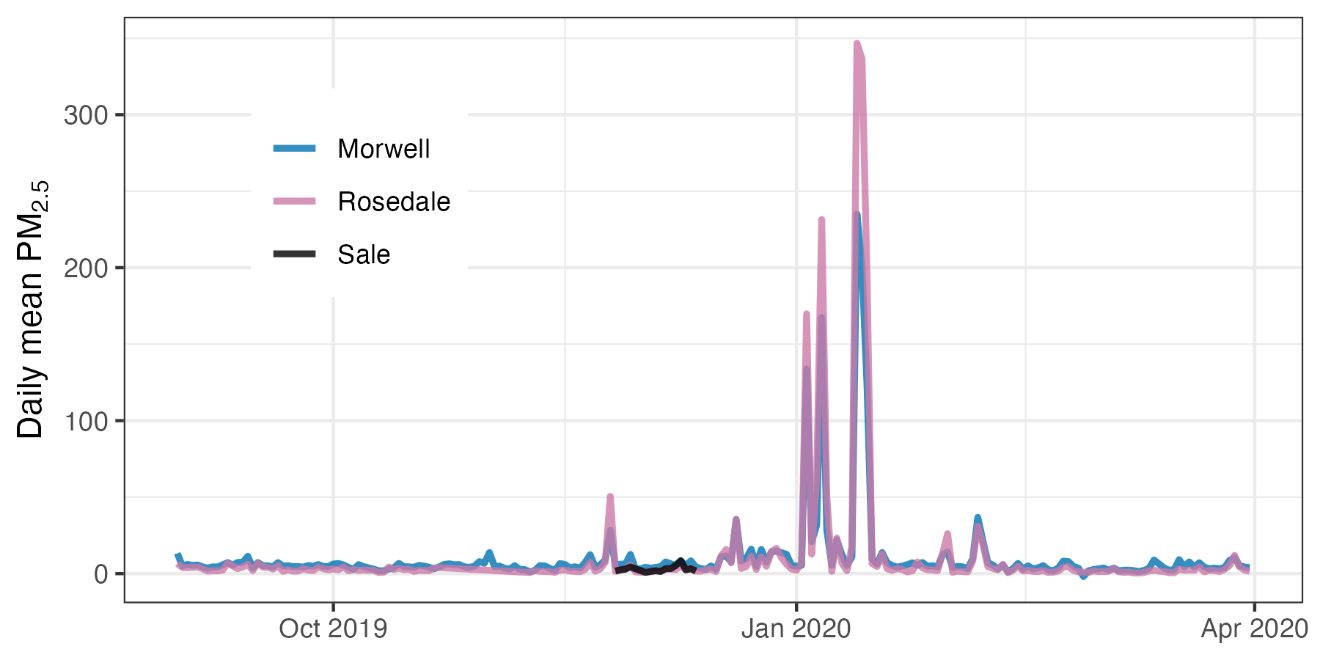

Figure S1. Completeness of PM_2.5_ monitoring station data in Morwell (exposure), Sale (control), and Rosedale (substitute for Sale) during the Black Summer.

A direct comparison between Morwell and Rosedale using quantile regression, determining whether there were differences at each percentile of their relative distributions, indicated background PM_2.5_ was slightly higher (~2.5µg/m^3^) in Morwell at nearly every quantile (Figure S2). Interestingly, the peaks at the 90^th^ percentile were not significantly different. While the confidence intervals for Rosedale become relatively large, indicating a lack of precision, the 90^th^ percentile points are nearly the same. Arguably, these higher quantiles matter most in terms of respiratory effects from smoke exposure during the Black Summer.

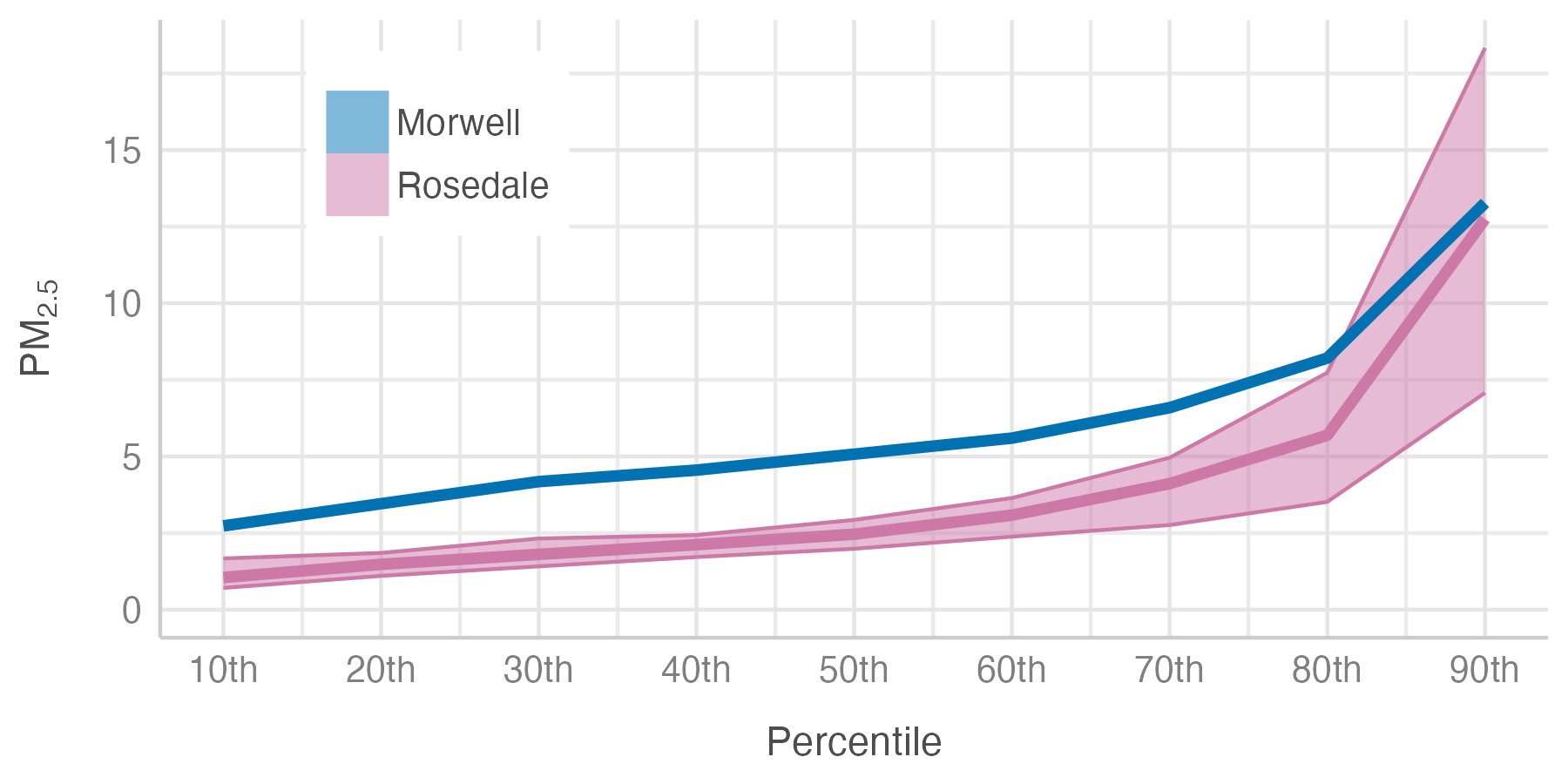

Figure S2. Quantile regression comparison of PM_2.5_ exposure during the Black Summer, between Morwell (exposed to mine fire smoke in 2014) and Rosedale (proxy for Sale, unexposed to the mine fire).

#### Further Supporting Information Tables and Figures

Table S1 Comparison of Round 1 characteristics according to participation status in Round 2.

| **R1 Characteristic** | **R2 non-participants** | **R2 Participants** |  |
| --- | --- | --- | --- |
|  | **N=190** | **N=329** | **P-value** |
| **Township, n (%)** |  |  |  |
| Sale | 61 (32%) | 112 (34%) | 0.672 |
| Morwell | 129 (68%) | 217 (66%) |  |
| **Female, n (%)** | 111 (58%) | 195 (59%) | 0.842 |
| **Education (Adult Survey), n (%)** |  |  |  |
| Secondary up to year 10 | 53 (28%) | 66 (20%) | 0.077 |
| Secondary year 11-12 | 40 (21%) | 63 (19%) |  |
| Certificate, University or other Tertiary Institute degree | 95 (51%) | 196 (60%) |  |
| **Employment status, n (%)** |  |  |  |
| Employed | 79 (42%) | 150 (46%) | 0.652 |
| Retired | 64 (34%) | 95 (29%) |  |
| Unemployed/unable to work | 25 (13%) | 49 (15%) |  |
| Other (home duties, study, other) | 22 (12%) | 35 (11%) |  |
| **Smoking status, n (%)** |  |  |  |
| Non-smoker | 87 (46%) | 162 (49%) | 0.042 |
| Ex-smoker | 62 (33%) | 124 (38%) |  |
| Current smoker | 41 (22%) | 43 (13%) |  |
| **Self-reported asthma, n (%)** | 75 (39%) | 154 (47%) | 0.113 |
| **Spirometric COPD, n (%)** | 24 (13%) | 35 (11%) | 0.417 |
| **Age, mean (SD)** | 55.6 (17.4) | 53.8 (15.2) | 0.247 |
| **Height (cm), mean (SD)** | 165.3 (9.7) | 166.8 (9.0) | 0.062 |
| **Weight (kg), mean (SD)** | 84.5 (22.8) | 86.2 (20.3) | 0.408 |
| **BMI (kg/m^2^), mean (SD)** | 30.9 (7.7) | 31.0 (7.273) | 0.920 |
| **At the time of the mine fire: Daily exposure to fire-related PM_2.5_ (µg/m^3^), median[Q1-Q3]** | 8.0 (0.0-14.0) | 7.0 (0.0-14.0) | 0.689 |
| **Cigarette pack years in smokers, median[Q1-Q3]** | 17.0 (7.0-36.0) | 15.0 (5.0-30.0) | 0.211 |

*Abbreviations*: R1 = Round 1, R2 = Round 2, SD = Standard Deviation, 95%CI = 95% Confidence Interval BMI=Body Mass Index; COPD: Chronic Obstructive Pulmonary Disease; PM_2·5_ = particulate matter <2·5 µg/m³, Q1 = first quartile, Q3 = third quartile.

*Missing*: education, *n* = 6; COPD, *n* = 13.

Table S2: Round 1 respiratory outcome means (z-scores or transformed) for R2 participants compared with non-participants.

| **Round 1 Outcome** | **R2 non-participants** | **R2 Participants** |  |
| --- | --- | --- | --- |
|  | **N=190** | **N=329** |  |
|  | **Mean (SD)** | **Mean (SD)** | **P-value** |
| **Spirometry (**z-scores) |  |  |  |
| Pre BD FEV_1_ | -0.53 (1.21) | -0.47 (1.16) | 0.609 |
| Pre BD FVC | -0.21 (1.05) | -0.13 (1.03) | 0.403 |
| Pre BD FEV_1_/FVC | -0.62 (1.16) | -0.60 (1.07) | 0.843 |
| Pre BD FEF_25-75%_ | -0.57 (1.21) | -0.56 (1.14) | 0.917 |
| Post BD FEV_1_ | -0.23 (1.23) | -0.15 (1.11) | 0.490 |
| Post BD FVC | -0.06 (1.01) | 0.03 (0.96) | 0.315 |
| Post BD FEV_1_/FVC | -0.35 (1.18) | -0.31 (1.09) | 0.720 |
| Post BD FEF_25-75%_ | -0.21 (1.26) | -0.15 (1.19) | 0.603 |
| **Gas transfer (**z-scores) |  |  |  |
| T_L_co | -0.42 (1.48) | -0.06 (1.25) | 0.006 |
| Hb corrected T_L_co | -0.37 (1.47) | -0.04 (1.21) | 0.011 |
| V_A_ | -0.48 (1.07) | -0.40 (1.02) | 0.399 |
| Kco | -0.03 (1.35) | 0.29 (1.23) | 0.008 |
| Hb corrected Kco | 0.02 (1.36) | 0.32 (1.20) | 0.017 |
| **Oscillometry** (non-linear transformed) |  |  |  |
| Baseline ln(R5) | 1.36 (0.41) | 1.33 (0.38) | 0.450 |
| Pre BD ln([R5-R19] +1) | 0.53 (0.43) | 0.48 (0.44) | 0.233 |
| Pre BD exp(X5) | 0.24 (0.16) | 0.26 (0.16) | 0.182 |
| Pre BD ln(Ax5) | 2.21 (0.97) | 2.04 (0.99) | 0.064 |
| Post BD ln(R5) | 1.27 (0.36) | 1.22 (0.36) | 0.144 |
| Post BD ln([R5-R19] +1) | 0.48 (0.39) | 0.41 (0.36) | 0.039 |
| Post BD exp(X5) | 0.28 (0.17) | 0.32 (0.17) | 0.027 |
| Post BD ln(Ax5) | 1.96 (0.94) | 1.74 (0.92) | 0.014 |

*Abbreviations*: R1 = Round 1, R2 = Round 2, SD = Standard Deviation, 95%CI = 95% Confidence Interval, FEV_1_ = Forced expiratory volume in 1 second, FVC = Forced vital capacity, FEF = Forced expiratory flow, BD = Bronchodilator, T_L_co = Transfer/diffusion factor of the lung for carbon monoxide, Hb = haemoglobin, V_A_ = Alveolar volume, Kco = Carbon monoxide transfer coefficient, R5 = resistance at 5Hz, R5-19 = difference in resistance at 5 and 19Hz, , X5 = reactance at 5Hz, Ax5 = area under the reactance curve at 5Hz

*Missing*: Oscillometry pre BD R1, n=44; Oscillometry post BD R1, n=41; Spirometry pre BD R1, n=11; Spirometry post BD R1, n=11; Gas transfer R1, n=11.

Table S3 Respiratory outcome crude mean scores (SD) by assessment round and mean differences for those completing both rounds (R2-R1).

| **Outcomes** | **Round 1** | **Round 1**  (subset who participated in R2) | **Round 2** | **Differences (R2-R1)** | | |
| --- | --- | --- | --- | --- | --- | --- |
|  | **N=519** | **N=329** | **N=329** |  | **(N=329)** |  |
|  | Mean (SD) | Mean (SD) | Mean (SD) | Mean difference | 95% CI | P-value |
| **Spirometry** |  |  |  |  |  |  |
| Pre BD FEV_1_ L | 2.76 (0.86) | 2.80 (0.84) | 2.77 (0.88) | -0.04 | (-0.07, -0.01) | 0.004 |
| Pre BD FVC L | 3.67 (1.04) | 3.73 (1.01) | 3.80 (1.06) | 0.06 | (0.03, 0.1) | <0.001 |
| Pre BD FEV_1_/FVC | 75.00 (9.66) | 75.14 (9.11) | 72.54 (9.06) | -2.57 | (-3.01, -2.13) | <0.001 |
| Pre BD FEF_25-75%_ L/s | 2.40 (1.16) | 2.42 (1.15) | 2.17 (1.12) | -0.25 | (-0.3, -0.2) | <0.001 |
| Post BD FEV_1_ L | 2.89 (0.87) | 2.94 (0.83) | 2.88 (0.89) | -0.07 | (-0.09, -0.04) | <0.001 |
| Post BD FVC L | 3.75 (1.01) | 3.81 (0.98) | 3.83 (1.05) | 0.01 | (-0.03, 0.04) | 0.703 |
| Post BD FEV_1_/FVC | 76.96 (9.88) | 77.18 (9.20) | 75.09 (9.73) | -2.03 | (-2.38, -1.68) | <0.001 |
| Post BD FEF_25-75%_ L/s | 2.73 (1.31) | 2.78 (1.29) | 2.54 (1.27) | -0.24 | (-0.29, -0.2) | <0.001 |
| **Gas transfer** |  |  |  |  |  |  |
| T_L_co mL/min/mmHg | 22.94 (7.04) | 23.64 (6.82) | 22.67 (6.75) | -0.97 | (-1.27, -0.66) | <0.001 |
| Hb corrected T_L_co | 23.02 (6.88) | 23.67 (6.59) | 22.58 (6.59) | -1.10 | (-1.4, -0.8) | <0.001 |
| V_A_ L | 5.07 (1.17) | 5.14 (1.16) | 5.07 (1.13) | -0.07 | (-0.11, -0.02) | 0.005 |
| Kco mL/min/mmHg/L | 4.52 (0.90) | 4.61 (0.85) | 4.46 (0.87) | -0.15 | (-0.2, -0.1) | <0.001 |
| Hb corrected Kco | 4.55 (0.88) | 4.62 (0.82) | 4.44 (0.85) | -0.18 | (-0.23, -0.13) | <0.001 |
| **Oscillometry** |  |  |  |  |  |  |
| Pre BD R5 cmH_2_O/L/sec | 4.12 (1.55) | 4.07 (1.56) | 4.08 (1.54) | -0.09 | (-0.23, 0.06) | 0.241 |
| Pre BD R5-R19 cmH_2_O/L/sec | 0.82 (0.84) | 0.79 (0.84) | 0.69 (0.84) | -0.16 | (-0.24, -0.07) | <0.001 |
| Pre BD X5 cmH_2_O/L/sec | -1.79 (1.23) | -1.73 (1.16) | -1.87 (1.36) | -0.04 | (-0.16, 0.07) | 0.481 |
| Pre BD Ax5 | 13.18 (14.68) | 12.49 (13.75) | 14.95 (18.08) | 1.05 | (-0.5, 2.6) | 0.182 |
| Post BD R5 cmH_2_O/L/sec | 3.68 (1.34) | 3.60 (1.28) | 3.60 (1.50) | -0.13 | (-0.26, -0.01) | 0.041 |
| Post BD R5-R19 cmH_2_O/L/sec | 0.66 (0.69) | 0.61 (0.64) | 0.56 (0.77) | -0.11 | (-0.18, -0.03) | 0.004 |
| Post BD X5 cmH_2_O/L/sec | -1.47 (1.00) | -1.38 (0.85) | -1.60 (1.19) | -0.08 | (-0.17, 0) | 0.051 |
| Post BD Ax5 | 9.62 (10.93) | 8.65 (9.21) | 10.78 (13.55) | 0.47 | (-0.52, 1.46) | 0.352 |

*Abbreviations:* R1 = Round 1, R2 = Round 2; SD = Standard Deviation, 95%CI = 95% Confidence Interval, FEV_1_ = Forced expiratory volume in 1 second, FVC = Forced vital capacity, FEF = Forced expiratory flow, BD = Bronchodilator, T_L_co = Transfer/diffusion factor of the lung for carbon monoxide, Hb = haemoglobin, V_A_ = Alveolar volume, Kco = Carbon monoxide transfer coefficient, R5 = resistance at 5Hz, R5-19 = difference in resistance at 5 and 19Hz, X5 = reactance at 5Hz, Ax5 = area under the reactance curve at 5Hz.

*Missing*: Oscillometry pre BD R1, n=44; Oscillometry post BD R1, n=41; Spirometry pre BD R1, n=11; Spirometry post BD R1, n=11; Gas transfer R1, n=11; Oscillometry pre BD R2, n=3; FOT post BD R2, n=2; Spirometry pre BD R2, n=6; Spirometry post BD R2, n=7; Gas transfer R2, n=4.

Table S4. Oscillometry mean z-scores (SD) by assessment round and mean differences (R2-R1).

| **Outcomes** | **Round 1** | **Round 1**  (subset who participated in R2) | **Round 2** | | **Differences (R2-R1)** | | |
| --- | --- | --- | --- | --- | --- | --- | --- |
|  | **N=519** | **N=329** | **N=329** | |  | **(N=329)** |  |
|  | Mean (SD) | Mean (SD) | Mean (SD) | | Mean difference | 95% CI | P-value |
| Pre BD R5 | 0.29 (1.23) | 0.25 (1.21) | | 0.21 (1.26) | -0.06 | (-0.18, 0.07) | 0.368 |
| Pre BD X5 | 0.62 (2.12) | 0.55 (2.00) | | 0.68 (2.22) | 0.02 | (-0.19, 0.23) | 0.869 |
| Pre BD Ax5 | 0.98 (1.25) | 0.93 (1.23) | | 0.95 (1.40) | -0.04 | (-0.17, 0.1) | 0.596 |
| Post BD R5 | -0.12 (1.20) | -0.16 (1.13) | | -0.28 (1.33) | -0.15 | (-0.27, -0.04) | 0.007 |
| Post BD X5 | -0.05 (1.81) | -0.19 (1.57) | | 0.11 (2.04) | 0.15 | (-0.02, 0.32) | 0.079 |
| Post BD Ax5 | 0.59 (1.20) | 0.51 (1.15) | | 0.50 (1.42) | -0.08 | (-0.19, 0.04) | 0.197 |

*Abbreviations*: SD = Standard Deviation, 95%CI = 95% Confidence Interval, BD = Bronchodilator, R5 = resistance at 5Hz, X5 = reactance at 5Hz, Ax5 = area under the reactance curve at 5Hz.

*Missing*: Oscillometry pre BD R1, n=44; Oscillometry post BD R1, n=41; Oscillometry pre BD R2, n=6; Oscillometry post BD R2, n=6.

Table S5. Spirometry test quality.

| **Round** | **Total tests available N** | **Included tests** | | **Excluded tests** |
| --- | --- | --- | --- | --- |
|  |  | **A or B grade*** | **Other than A or B grade*** |  |
|  | N | n (%) | n (%) | n (%) |
| R1 Pre BD | 513 | 476 | 32 | 5 |
| R1 Post BD | 505 | 480 | 22 | 3 |
| R2 Pre BD | 328 | 312 | 11 | 5 |
| R2 Post BD | 327 | 317 | 5 | 5 |

* Graded as per ATS/ERS spirometry criteria at time of assessment

*Abbreviations:* R1 = Round 1, R2 = Round 2; BD = Bronchodilator.

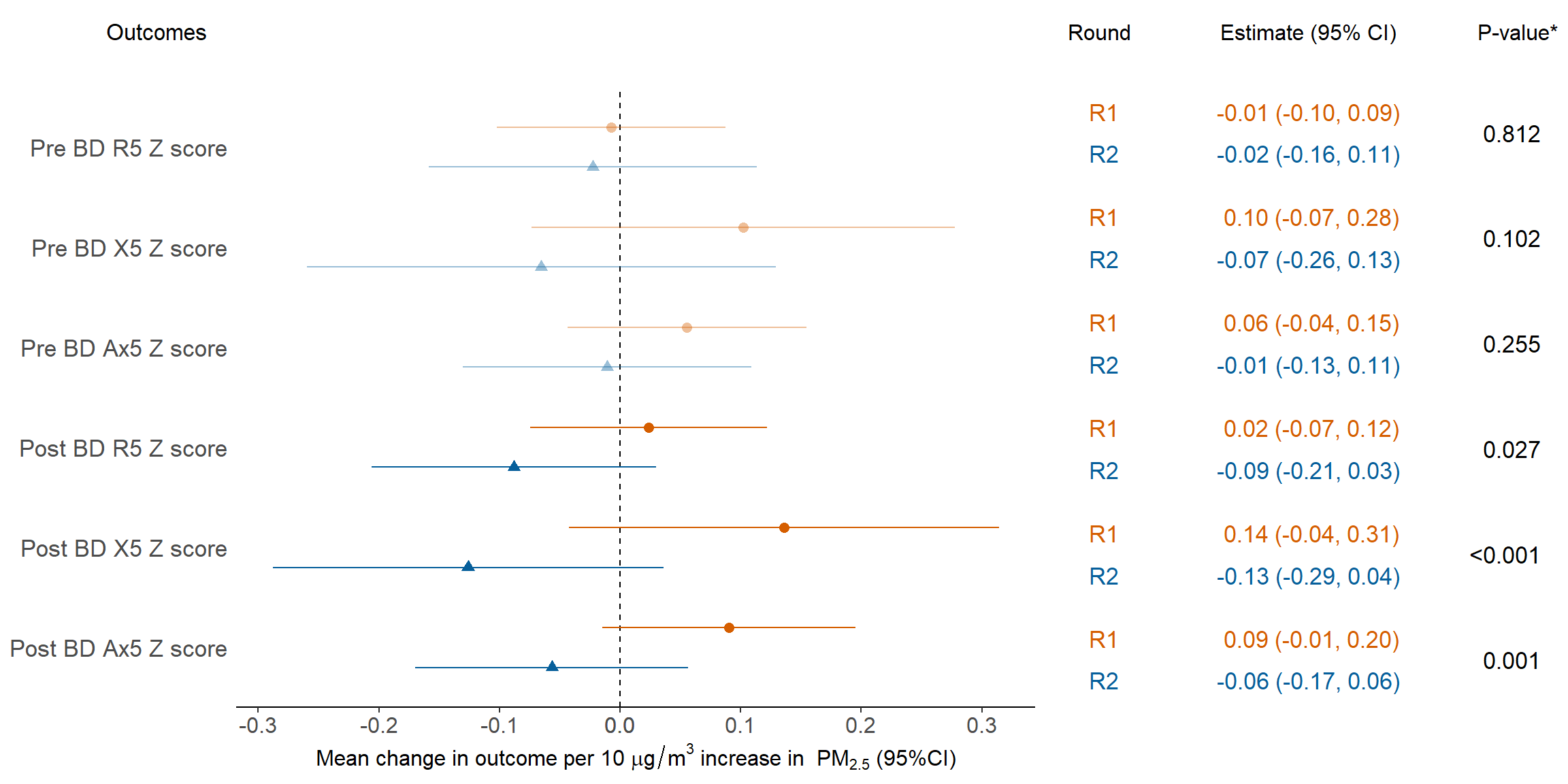

Figure S4: Models of Oscillometry Z Scores as a function of mine fire-related PM_2.5._

*Abbreviations*: R1 = Round 1, R2 = Round 2, SD = Standard Deviation, 95%CI = 95% Confidence Interval, PM_2·5_ = particulate matter <2·5 µg/m³, R5 = resistance at 5Hz, R5-19 = difference in resistance at 5 and 19Hz, X5 = reactance at 5Hz, Ax5 = area under the reactance curve at 5Hz.

* All models were adjusted for education, employment, asthma, spirometric COPD and smoking status. Pre BD outcome also adjusted for whether inhaled medication was withheld prior to spirometry.
